# Exploring causal relationships between COVID-19 and cardiometabolic disorders: A bi-directional Mendelian randomization study

**DOI:** 10.1101/2021.03.20.21254008

**Authors:** Yong Xiang, Carlos Kwan-Long Chau, Jinghong Qiu, Shitao Rao, Hon-Cheong So

**Author notes:** **Correspondence to: Hon-Cheong So,** Lo Kwee-Seong Integrated Biomedical Sciences Building, The Chinese University of Hong Kong, Shatin, Hong Kong.

## Abstract

**Background:** More than 100 million cases of COVID-19 have been reported worldwide. A number of risk factors for infection or severe infection have been identified, however observational studies were subject to confounding bias. In addition, there is still limited knowledge about the complications or medical consequences of the disease.

**Methods:** Here we performed bi-directional Mendelian randomization (MR) analysis to evaluate causal relationships between liability to COVID-19 (and severe/critical infection) and a wide range of around 30 cardiometabolic disorders (CMD) or traits. Genetic correlation (rg) was assessed by LD score regression(LDSC). The latest GWAS summary statistics from the COVID-19 Host Genetics Initiative was used, which comprised comparisons of general population controls with critically ill, hospitalized and any infected cases.

**Results:** Overall we observed evidence that liability to COVID-19 or severe infection may be causally associated with higher risks of type 2 diabetes mellitus(T2DM), chronic kidney disease(CKD), ischemic stroke (especially large artery stroke[LAS]) and heart failure(HF) when compared to the general population. On the other hand, our findings suggested that liability to atrial fibrillation (AF), stroke (especially LAS), obesity, diabetes (T1DM and T2DM), low insulin sensitivity and impaired renal function (low eGFR and diabetic kidney disease) may be causal risk factors for COVID-19 or severe disease. In genetic correlation analysis, T2DM, CAD, obesity, fasting insulin, CKD, gout, stroke and urate showed positive rg with critical or hospitalized infection. All above findings passed multiple testing correction at a false discovery rate (FDR)<0.05.

**Conclusions:** In summary, this study provides evidence for tentative bi-directional causal associations between liability to COVID-19 and severe disease and a number of CM disorders. Further replications and prospective studies are required to verify the findings.

## Introduction

More than 100 million cases of COVID-19 has been reported worldwide, and the pandemic has resulted in >2.4 million fatalities as at 17 Feb 2020. A number of risk factors for infection or severe infection have been identified, such as age, sex (male), obesity, diabetes mellitus, renal impairment, multi-comorbidities, among others ^1-5^. However, an important limitation is that most evidence came from observational studies, which were subject to risk of (residual) confounding, making it difficult to infer causality. In addition, there is still limited knowledge about the complications and medium- or long-term consequences of the disease. Similar to the study of risk factors, observational studies for disease consequences may be subject to confounding, such as confounding by the use of medications. Reverse causality is also possible. For example, a patient with a pre-existing but undiagnosed condition may be more prone to the infection, but the condition may only become diagnosed after check-ups in hospital. The condition may therefore be mistaken as a consequence of the infection.

Here we employed Mendelian randomization (MR) to explore potential causal relationships between COVID-19 and cardiometabolic disorders (CMD), including the effect of COVID-19 on CMD and CMD as risk factors for COVID-19. Intuitively, MR studies the *genetically predicted exposure* and its association with the outcome, and is much less susceptible to confounding and reverse causality. MR can therefore shed light on potential *causal relationships* between the exposure and the outcome. By studying genetic predisposition to COVID-19 or CMD as exposure, there is much lower risk that the studied effects are due to confounding factors alone.

We focused on cardiometabolic abnormalities and disorders here as a number of observational studies have shown such disorders may increase the risk or severity of infection ^6,7^. Although less widely studied, several studies also raised the possibility that COVID-19 may be associated with cardiovascular consequences ^8-11^, either in the short term or over longer periods of time. For example, it has been suggested that COVID-19 infection may be associated with increased risks of myocarditis, arrhythmias, heart failure and venous thromboembolism. In a German study of 100 subjects ^12^who have recently recovered from the disease, 78% had abnormal cardiovascular magnetic resonance (CMR) findings, and 60% had evidence of ongoing inflammation. Interestingly, the risks of cardiac abnormalities were independent of underlying comorbidities or severity of infection. Another similar study on athletes with mild or asymptomatic infections showed CMR evidence of myocarditis or prior myocardial injury in ∼15% and 30.8% of patients respectively ^13^. While the clinical importance of these findings is uncertain and the long-term consequences cannot be derived from the above studies, they showed that cardiovascular consequences are possible among those recovered from the infection.

The link between COVID-19 infection and diabetes is another topic of high interest. Diabetes has been shown in numerous studies as a risk factor for susceptibility to and severity of infection ^14,15^. On the other hand, studies have raised the possibility of new-onset diabetes among infected patients ^16-19^. A recent systematic review and meta-analysis^19^ aggregated data over 8 studies and estimated a pooled proportion of 14.4% (95% CI: 5.9%-25.8%) of newly diagnosed diabetes in hospitalized COVID-19 patients. It remains to be investigated whether the phenomenon is temporary or long-lasting ^16,20^

While long-term follow-up (FU) for COVID-19 is not possible yet as it is a new disease, a study on hospitalized pneumonia patients showed elevated risk of cardiovascular diseases in both short-term (30 days to 1 year after hospitalization) and long term (up to 10 years), although the elevated risk was more marked within the 1st year of illness ^21^.

The above studies suggested possible associations between CM disorders and COVID-19, but as mentioned earlier, confounding and reverse causality may render causal interpretations difficult. Here we conducted a comprehensive study using a variety of MR methods and settings, with inclusion of a wide range of CM disorders, and employed the latest GWAS results from the COVID-19 Host Genetics Initiative (HGI). As a novel aspect of this study, we not only investigated CMD as risk factors as in previous studies (e.g. ^22^), but also investigated how liability to COVID-19 may causally increase the risk of CMD.

## Methods

### GWAS data

#### COVID-19 data

We extracted GWAS summary statistics from the COVID-19 Host Genetics Initiative, release 5 (updated Jan 18 2021). For details please refer to the website ^23^and ^24^. The latest GWAS analysis methodology is available at https://docs.google.com/document/d/16ethjgi4MzlQeO0KAW_yDYyUHdB9kKbtfuGW4XYVKQg/edit.

We mainly considered three sets of summary statistics which compared very severe/critically ill, hospitalized, and any COVID-19 cases against unscreened population controls (analysis A2, B2, C2 respectively). The detailed definitions of the COVID-19 phenotypes are available at https://docs.google.com/document/d/1okamrqYmJfa35ClLvCt_vEe4PkvrTwggHq7T3jbeyCI/edit. Briefly, very severe or critically ill cases are defined as hospitalized laboratory-confirmed cases who required respiratory support or with cause of mortality related to the infection. “Hospitalized cases” are defined as those with laboratory-confirmed infection and hospitalized due to related symptoms. For analysis C2, “infected cases” included those with laboratory-confirmed infection, physician or electronic health record-confirmed infection or self-reported positive cases. These three datasets were chosen mainly because the sample sizes were among the largest which improves the power for MR analysis (A2: 5870 cases/1,155,203 controls; B2, 11,829 cases/1,725,210 controls; C2: 42,557 cases/1,424,707 controls). We chose the summary results excluding UK Biobank study to avoid sample overlap with GWAS results of cardiometabolic traits which often included UKBB subjects.

#### Cardiometabolic disorders

The list of cardiometabolic (CM) disorders under study is presented in Table 1. In brief, we included mainly disease traits including atrial fibrillation (AF), coronary artery disease (CAD), heart failure (HF), stroke (including different subtypes), venous thromboembolism (VTE), Type 1 diabetes mellitus (T1DM), Type 2 diabetes mellitus (T2DM), chronic kidney disease (CKD) including diabetic kidney disease (DKD) [and related measures such as estimated glomerular filtration rate (eGFR), blood urea nitrogen (BUN), Urine Albumin-to-Creatinine Ratio (UACR)], gout, and several measures related to glycemic control or insulin resistance. We are including a number of diabetes-related phenotypes as DM is established to be associated with COVID-19 infection and severity{Lim, 2021 #72}, and our previous MR study of pulmonary ACE2 expression{Rao, 2020 #73} also suggested DM and related traits may be a causal risk factors for infection. The majority of the samples are European in ancestry but we also included meta-analysis of trans-ethnic samples for larger sample sizes. All GWAS summary statistics were corrected for population stratification.

**Table 1.** Overview of disorders/traits included in MR analysis

| Category | Traits | PMID or reference | Sample size, N | Ancestry of participants |
| --- | --- | --- | --- | --- |
| COVID-19 | A2 (Very severe/critical covid vs. population) | 32404885 | 1161073 | Mostly European |
|  | B2 (Hospitalized covid vs population) | 32404885 | 1737039 | Mostly European |
|  | C2 (covid vs population) | 32404885 | 1467264 | Mostly European |
| AF | Atrial Fibrillation | 30061737 | 1030836 | European |
|  | Atrial Fibrillation | 29892015 | 588190 | Multi-Ethnic |
| CAD | Coronary Artery Disease | 28714975 | 148172 | European |
| Heart Failure | Heart Failure (HF) | 31919418 | 977323 | Mostly European |
| Obesity | Obesity | 23563607 | 344311 | European |
| Stroke | Any Stroke (AS) | 29531354 | 446696 | European |
|  | Any Stroke | 29531354 | 521612 | Multi-Ethnic |
|  | Any Ischemic Stroke (AIS) | 29531354 | 440328 | European |
|  | Any Ischemic Stroke | 29531354 | 514791 | Multi-Ethnic |
|  | Cardioembolic Stroke (CES) | 29531354 | 413304 | European |
|  | Cardioembolic Stroke | 29531354 | 463456 | Multi-Ethnic |
|  | Large Artery Stroke (LAS) | 29531354 | 410484 | European |
|  | Large Artery Stroke | 29531354 | 461138 | Multi-Ethnic |
|  | Small Vessel Stroke (SVS) | 29531354 | 411497 | European |
|  | Small Vessel Stroke | 29531354 | 466160 | Multi-Ethnic |
| CKD-related | Blood Urea Nitrogen (BUN) | 31152163 | 243029 | European |
|  | Blood Urea Nitrogen | 31152163 | □416178 | Multi-Ethnic |
|  | Chronic Kidney Disease (CKD) | 31152163 | 480698 | European |
|  | Chronic Kidney Disease | 31152163 | 625219 | Multi-Ethnic |
|  | eGFR | 31152163 | 567460 | European |
|  | eGFR | 31152163 | 765348 | Multi-Ethnic |
|  | Microalbuminuria | 31511532 | 348954 | Multi-Ethnic |
|  | Gout | 31578528 | 763813 | Multi-Ethnic |
|  | UACR Diabetes mellitus | 31511532 | 554156 | Multi-Ethnic |
|  | UACR | 31511532 | 547361 | European |
|  | UACR | 31511532 | 564257 | Multi-Ethnic |
|  | Urate | 31578528 | 288649 | European |
|  | Urate | 31578528 | 457690 | Multi-Ethnic |
| Glycemic/DKD | Fasting Glucose (FG) | 33402679 | 151188 | European |
|  | Change in Fasting Glucose Over Time | 31263163 | 13807 | Multi-Ethnic |
|  | Fasting Insulin (FI) | 33402679 | 105056 | European |
|  | HbA1c | 28898252 | 159940 | European |
|  | Insulin Sensitivity Index (ISI) Model 1 | 27416945 | 16753 | European |
|  | Insulin Sensitivity Index (ISI) Model 2 | 27416945 | 16735 | European |
|  | Insulin Sensitivity Index (ISI) Model 3 | 27416945 | 16735 | European |
|  | T1D_ALL_DKD_EUR | 27647854 | 12251 | European |
|  | T1D_Early_DKD_EUR | 27647854 | 5454 | European |
|  | T1D_eGFR_EUR | 27647854 | 6335 | European |
|  | T1D_Late_DKD_EUR | 27647854 | 6878 | European |
|  | T2D_eGFR_EUR | 29703844 | 7056 | European |
| T1DM | Type 1 Diabetes | 32005708 | 38712 | European |
|  | Type 1 Diabetes | Pan-UKB team | 412700 | Multi-Ethnic |
| T2DM | Type 2 Diabetes | 30297969 | 898130 | European |
| VTE | Venous Thromboembolism | 31676865 | 223597 | Multi-Ethnic |

Note: PMID: PubMed identifier; UACR: Urinary Albumin-to-creatinine Ratio; eGFR: estimated glomerular filtration rate; ISI, modified Stumvoll Insulin Sensitivity Index (Model 1 was adjusted for age and sex; Model 2 was adjusted for age, sex, and body mass index; and Model 3 analysed the combined influence of the genotype effect adjusted for BMI and the interaction effect between the genotype and BMI on ISI); DKD, diabetic kidney disease.

### Mendelian randomization methods

Here we performed two-sample MR, in which the instrument-exposure and instrument-outcome associations were estimated in different samples.

We conducted MR with several different MR approaches based on different assumptions, including (1) ‘inverse-variance weighted’ (MR-IVW)^25^ ; (2) Egger regression (MR-Egger)^26^; (3) weighted median (WM); (4) GSMR; and (5) MR-RAPS. Multiple testing due to use of different methods was accounted for by false discovery rate (FDR) correction.

The IVW approach is one of the most widely employed approaches for MR which assumes balanced pleiotropic effects. Horizontal pleiotropy is one of the concerns of MR, in which the genetic instruments have effects on the outcome other than through effects on the exposure. The Egger regression approach allows imbalanced pleiotropy and can produce valid estimate of the causal effects in such cases. MR-Egger (and also IVW) requires ^27^ the InSIDE assumption, i.e. the SNP-exposure effects are independent of the horizontal pleiotropic effects. The weighted median approach assumes at least half of the instruments are valid, and employs a median estimator to avoid influence by outliers. We employed the “TwoSampleMR” R package for the above methods. The method GSMR also takes into account of horizontal pleiotropy; it operates based on the exclusion of ‘outlier’ or heterogeneous genetic instruments that are likely pleiotropic (known as the ‘HEIDI-outlier’ method)^28^. The GSMR framework also employed a slightly different formula from the conventional IVW approach by modelling variance of both SNP-exposure and SNP-outcome coefficients. Correlated variants can be accommodated. We employed the GSMR R package from http://cnsgenomics.com/software/gsmr/. Please refer to ref^28^ for details.

The robust adjusted profile score (MR-RAPS) ^29^ is a recently developed approach that also considers the measurement error in SNP-exposure effects and was shown to be unbiased in the presence of many weak instruments. MR-RAPS allows both systematic and idiosyncratic pleiotropy. In other words, most pleiotropic effects are assumed to be distributed normally (with mean zero) but a few large pleiotropic effects can be present. For details please refer to ^30^. The “mr-raps” R package was used with default settings, allowing for overdispersion and shrinkage estimates.

#### Inclusion of a larger number of SNPs as instruments

One main difficulty of MR analysis here, especially for the study of causal effects of COVID-19 on other traits, is that the number of SNP instruments passing genome-wide significance is in general small, which limits the power of detecting causal associations. Small number of instruments also makes the assessment of horizontal pleiotropy difficult. Conventionally, only SNPs with genome-wide significance were included in MR. However, recent studies have proposed the use of a large number of ‘weak instruments’ ^30^.

A main concern of using a liberal p-value threshold is weak instrument bias, which tends to bias towards *the null* in two-sample MR ^31^. Since this bias is *conservative*, type I error control should not be affected. This is supported by several previous studies. Simulation studies by Wang et al. showed that that type I error control (for the null hypothesis of causal effect=0) is maintained for very weak instruments with F-statistics down to ∼1 (see Figure 1^32^). Another study^30^ also showed largely maintained type I error rates in simulations, when MR is conducted with liberal inclusion of variants (no. of instruments ∼900) not passing genome-wide significance.

To further verify that type I error (false positive rate) rate is maintained with a more liberal p-value threshold, we also performed additional simulation studies, based on the code and simulation method described in Qi et al. ^33^. We conducted simulations under two sample sizes (*N*=50,000 or 100,000 for exposure trait and halved for outcome), several liberal p-value thresholds (p=1e-4, 1e-3, 1e-2) and different proportion of invalid instruments (10% or 30%) with horizontal pleiotropy, under zero causal effect. We performed 1000 replications; balanced pleiotropy and InSIDE assumption were assumed. The five MR methods employed were tested. We observed that type I error rate were generally maintained (at ∼5% when p<0.05 regarded as significant) even when instruments were selected at liberal p-value levels (see Results and Table S1).

On the other hand, the causal effect estimate based on weak instruments tends to bias towards zero with many MR methods, which should be noted in the interpretation of results. To ameliorate this issue, the MR-RAPS approach can correct for such bias and the authors ^29^ showed valid estimation of causal effects up to a p-value threshold of 0.01. In another recent work ^30^, it was proposed that a ‘genome-wide’ MR design (the authors used ∼1100 top SNPs in their working examples) with MR-RAPS can improve statistical power and provide unbiased estimates of causal effects despite weak instruments. The same study and previous works ^34^ also showed MR-Egger with SIMEX correction could (partially) reduce the conservative bias of effect estimates, which we also employed in this study.

Based on the above studies and recent advances, here we adopted a more complex design considering more liberal p-value thresholds (pthres) for instruments to improve power. To avoid the arbitrariness of considering only one or two thresholds, we considered a range of thresholds [p=5e-8, 1e-7, 1e-6, 1e-5, 1e-4, 1e-3, 1e-2] and accounted for multiple testing by FDR.

#### Modelling correlated SNPs

Independent SNPs (r^2^ clumping threshold=0.001) were used as input for all methods. For IVW and GSMR, methodologies have been developed to handle correlated SNPs as well, which can improve the power of detecting causal associations. However, as raised by ^35^, if the SNPs are too highly correlated, the resulting estimates may be unstable and subject to inflation of type I error. This is because the SNP correlation matrix may be near-singular, which can also occur when a large number of SNPs are modelled due to haplotypes or linear combination of SNPs being correlated with other SNPs. Simulation studies by Burgess et al. ^35^ showed adequate type I error control at moderate correlations (rho) up to 0.4 to 0.6 with ∼320 variants. Based on the simulation findings, to prevent unstable estimates, for correlated-SNP analyses we set an r^2^ threshold of 0.2 and a threshold for the number of SNPs at 350. To avoid arbitrariness of setting a single LD-clumping cut-off, we performed MR analysis with correlated SNPs at four levels of r^2^ (0.05, 0.1, 0.15 and 0.2) and assessed the consistency of results, with correction of multiple testing by FDR ^35^. With the above settings, we did not observe abnormally low p-values or small SE signifying near-singularity of genetic correlation matrix. We employed the R packages “MendelianRandomization” and “gmsr” for MR-IVW and GSMR of correlated SNPs respectively.

Here we briefly described the IVW approach that is able to account for SNP correlations as described in Burgess et al.^36^. Briefly, assume 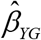 to be the vector of estimated regression coefficients when the outcome is regressed on genetic instruments and *σ*_*YG*_ to be the corresponding standard errors (SE), and 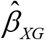 to be the estimated coefficients when the risk factor is regressed on the genetic instruments with SE *σ* _*XG*_. We also assume the correlation between two genetic variants G_1_ and G_2_ to be *ρ*_*G*1*G* 2_, and 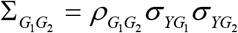.

The estimate from a weighted generalized linear regression can be formulated by

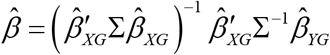

with SE

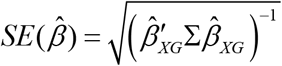

GSMR can also handle correlated SNPs based on a similar principle. Please refer to ^28^ for technical details.

#### Interpretation of MR causal estimate for binary exposure

For exposures that are binary (e.g. disease traits), the MR causal estimate should be regarded as the change in the outcome per log-odds change of the exposure. In other words, the causal estimate reflect the change of outcome for every 2.72-fold increase in the odds of the exposure. Assuming the exposure is not common, it is roughly equivalent to 2.72-fold increase in the *probability* of exposure trait, e.g. a change of disease/infection risk from 1% to 2.72%. Alternatively, we may consider the exposure as “disease liability” under a liability threshold model (measured on logit scale); the purpose is to evaluate the effect on outcome per unit increase in liability ^37^.

#### Steiger test of directionality

In brief, this test examines whether the instrument SNPs explain more variance for exposure than for the outcome ^38^. This serves to further confirm whether the causal direction is correct. We employed the mr_steiger function in “TwoSampleMR” for this test. We filtered away results whose causal direction is indicated as “FALSE” by the function.

#### Multiple testing control by FDR

Multiple testing was controlled by the false discovery rate (FDR) approach according to the Benjamini-Hochberg method^39^, which controls the expected *proportion* of false positives among the rejected hypotheses. In this study we set a FDR threshold of 0.05 to declare significance. FDR calculation was stratified by each combination of MR method and CM trait. Note that FDR control by the Benjamini-Hochberg method is also valid under positive (regression) dependency of hypothesis tests^40^.

#### Genetic correlation analysis

Genetic correlation analysis with the LD score regression program ^41^ following default settings. The method evaluates genetic overlap between pairs of disorders, although it is not designed for inferring causality.

## Results

### Simulation results

Full results are shown in Table S1. We observed adequate type I error control (∼5% when p<0.05 regarded as significant) across most scenarios at the three liberal p-value thresholds (pthres=1e-4, 1e-3, 1e-2). The results were similar with different samples sizes and proportion of invalid instruments. There was a slight increase in type I error (up to ∼8%) for MR-Egger at pthres=1e-2 when the proportion of invalid of instruments was high (30%); for GSMR, there was increase in type I error when *N*=100,000 at pthres=1e-4 with 30% invalid instruments. (GSMR adopts an ‘outlier-removal’ approach and therefore may require a larger proportion of valid instruments). However, in almost all other scenarios type I errors were maintained, and in this study we considered evidence across different methods instead of focusing a single approach. Overall, in addition to the previous literature, the results provides further support for the use of a larger number of less significant SNPs as instruments.

### MR results: liability to COVID-19 as exposure and cardiometabolic disorders as outcome

Here we highlight results which survive multiple testing correction at FDR<0.05. Results are presented in Tables 2-4 and S2. If significant results are observed across multiple pthres and/or r^2^-clumping thresholds, for simplicity we would present the one with the most stringent pthres and lowest r^2^ in main tables.

**Table 2.** MR results with liability to COVID-19 as exposure and cardiometabolic disorders (except Type 2 DM) as outcome (independent SNPs as instruments)

| exposure | outcome | method | nsnps | b | se | OR | lowerCI_OR | upperCI_OR | p_thres | pval | p.adjust |
| --- | --- | --- | --- | --- | --- | --- | --- | --- | --- | --- | --- |
| C2 | AF_TRANS | GSMR | 27 | 0.095 | 0.029 | 1.100 | 1.040 | 1.163 | 1.00E-05 | 8.93E-04 | 1.52E-02 |
| B2 | CES_EUR | MR Egger | 428 | 0.115 | 0.042 | 1.122 | 1.033 | 1.219 | 1.00E-03 | 6.67E-03 | 4.23E-02 |
| C2 | CES_EUR | MR Egger | 466 | 0.242 | 0.066 | 1.274 | 1.119 | 1.450 | 1.00E-03 | 2.87E-04 | 2.73E-03 |
| B2 | CES_TRANS | MR Egger | 424 | 0.120 | 0.041 | 1.127 | 1.040 | 1.222 | 1.00E-03 | 3.75E-03 | 2.38E-02 |
| C2 | CES_TRANS | MR Egger | 457 | 0.211 | 0.063 | 1.235 | 1.091 | 1.398 | 1.00E-03 | 9.04E-04 | 8.59E-03 |
| B2 | CKD_EUR | GSMR | 427 | 0.021 | 0.006 | 1.021 | 1.009 | 1.035 | 1.00E-03 | 1.10E-03 | 9.35E-03 |
| B2 | CKD_EUR | SIMEX | 427 | 0.025 | 0.007 | 1.026 | 1.011 | 1.040 | 1.00E-03 | 5.39E-04 | 6.46E-03 |
| B2 | CKD_TRANS | GSMR | 430 | 0.026 | 0.006 | 1.026 | 1.015 | 1.037 | 1.00E-03 | 3.61E-06 | 3.07E-05 |
| B2 | CKD_TRANS | IVW | 430 | 0.029 | 0.006 | 1.030 | 1.018 | 1.041 | 1.00E-03 | 3.50E-07 | 1.40E-06 |
| B2 | CKD_TRANS | MR Egger | 430 | 0.068 | 0.016 | 1.071 | 1.037 | 1.106 | 1.00E-03 | 3.59E-05 | 1.44E-04 |
| B2 | CKD_TRANS | MR-RAPS | 430 | 0.033 | 0.006 | 1.034 | 1.021 | 1.046 | 1.00E-03 | 9.79E-08 | 3.91E-07 |
| B2 | CKD_TRANS | SIMEX | 430 | 0.031 | 0.006 | 1.031 | 1.019 | 1.043 | 1.00E-03 | 8.54E-07 | 1.02E-05 |
| B2 | CKD_TRANS | Wt_median | 430 | 0.025 | 0.008 | 1.026 | 1.010 | 1.042 | 1.00E-03 | 1.31E-03 | 5.22E-03 |
| C2 | FI_EUR | IVW | 113 | -0.024 | 0.008 | 0.977 | 0.962 | 0.992 | 1.00E-04 | 3.03E-03 | 4.24E-02 |
| C2 | FI_EUR | MR Egger | 390 | -0.049 | 0.016 | 0.952 | 0.922 | 0.984 | 1.00E-03 | 3.11E-03 | 4.35E-02 |
| C2 | FI_EUR | SIMEX | 113 | -0.025 | 0.008 | 0.975 | 0.960 | 0.990 | 1.00E-04 | 1.56E-03 | 3.75E-02 |
| B2 | HeartFailure | IVW | 846 | 0.014 | 0.004 | 1.014 | 1.005 | 1.023 | 1.00E-02 | 1.93E-03 | 3.08E-02 |
| B2 | HeartFailure | MR Egger | 422 | 0.045 | 0.016 | 1.046 | 1.014 | 1.079 | 1.00E-03 | 5.38E-03 | 4.30E-02 |
| B2 | HeartFailure | MR-RAPS | 846 | 0.015 | 0.005 | 1.015 | 1.005 | 1.025 | 1.00E-02 | 2.36E-03 | 3.78E-02 |
| B2 | HeartFailure | SIMEX | 846 | 0.015 | 0.005 | 1.015 | 1.006 | 1.025 | 1.00E-02 | 1.81E-03 | 4.35E-02 |
| B2 | LAS_EUR | IVW | 430 | 0.056 | 0.018 | 1.057 | 1.020 | 1.096 | 1.00E-03 | 2.33E-03 | 2.21E-02 |
| B2 | LAS_EUR | MR Egger | 430 | 0.173 | 0.052 | 1.189 | 1.073 | 1.316 | 1.00E-03 | 9.95E-04 | 9.45E-03 |
| B2 | LAS_EUR | MR-RAPS | 430 | 0.061 | 0.020 | 1.063 | 1.022 | 1.105 | 1.00E-03 | 2.23E-03 | 2.12E-02 |
| B2 | LAS_EUR | SIMEX | 430 | 0.058 | 0.020 | 1.060 | 1.020 | 1.101 | 1.00E-03 | 3.13E-03 | 3.75E-02 |
| C2 | LAS_EUR | MR Egger | 921 | 0.215 | 0.069 | 1.240 | 1.084 | 1.419 | 1.00E-02 | 1.76E-03 | 1.11E-02 |
| B2 | LAS_TRANS | MR Egger | 844 | 0.107 | 0.034 | 1.113 | 1.041 | 1.190 | 1.00E-02 | 1.76E-03 | 3.35E-02 |
| B2 | SVS_EUR | MR Egger | 851 | 0.108 | 0.036 | 1.114 | 1.038 | 1.197 | 1.00E-02 | 3.00E-03 | 2.85E-02 |
| C2 | SVS_EUR | MR Egger | 920 | 0.192 | 0.061 | 1.212 | 1.074 | 1.367 | 1.00E-02 | 1.84E-03 | 2.85E-02 |
| A2 | T2D_eGFR_EUR | MR Egger | 141 | -3.869 | 1.102 | 0.021 | 0.002 | 0.181 | 1.00E-04 | 6.00E-04 | 1.14E-02 |

LD-clumping was performed at $r^2=0.001$ .
Nsnps: number of SNPs; b, causal effect estimate; se, standard error; OR, odds ratio (for every log odds increase in exposure risk); lowerCI\_OR and upperCI\_OR, lower and upper CI of OR; p\_thres, p-value threshold for inclusion as instrument; pval: p-value for MR analysis; p.adjust, FDR(false discovery rate)-adjusted p-value.

**Table 3.** MR results with liability to COVID-19 as exposure and Type 2 DM as outcome (independent SNPs as instruments)

| exposure | outcome | method | nsnps | b | se | OR | lowerCI_OR | upperCI_OR | pval | p_thres | p.adjust |
| --- | --- | --- | --- | --- | --- | --- | --- | --- | --- | --- | --- |
| A2 | T2DM | GSMR | 6 | 0.033 | 0.015 | 1.034 | 1.004 | 1.064 | 2.41E-02 | 1.00E-07 | 4.55E-02 |
| A2 | T2DM | IVW | 6 | 0.034 | 0.015 | 1.035 | 1.006 | 1.065 | 1.94E-02 | 5.00E-08 | 4.10E-02 |
| A2 | T2DM | MR-RAPS | 6 | 0.037 | 0.015 | 1.038 | 1.007 | 1.069 | 1.58E-02 | 5.00E-08 | 3.76E-02 |
| A2 | T2DM | SIMEX | 143 | 0.020 | 0.007 | 1.020 | 1.007 | 1.033 | 3.47E-03 | 1.00E-04 | 2.23E-02 |
| A2 | T2DM | Wt_median | 143 | 0.022 | 0.007 | 1.022 | 1.009 | 1.036 | 1.27E-03 | 1.00E-04 | 2.41E-02 |
| B2 | T2DM | GSMR | 33 | 0.032 | 0.012 | 1.033 | 1.009 | 1.057 | 7.36E-03 | 1.00E-05 | 2.50E-02 |
| B2 | T2DM | IVW | 34 | 0.044 | 0.017 | 1.045 | 1.009 | 1.081 | 1.25E-02 | 1.00E-05 | 3.86E-02 |
| B2 | T2DM | MR-RAPS | 34 | 0.043 | 0.017 | 1.044 | 1.010 | 1.080 | 1.02E-02 | 1.00E-05 | 3.24E-02 |
| B2 | T2DM | SIMEX | 143 | 0.025 | 0.009 | 1.025 | 1.007 | 1.044 | 8.17E-03 | 1.00E-04 | 3.92E-02 |
| C2 | T2DM | GSMR | 463 | 0.020 | 0.008 | 1.020 | 1.005 | 1.036 | 1.15E-02 | 1.00E-03 | 2.80E-02 |
| C2 | T2DM | IVW | 28 | 0.085 | 0.035 | 1.089 | 1.017 | 1.167 | 1.50E-02 | 1.00E-05 | 3.86E-02 |
| C2 | T2DM | MR-RAPS | 28 | 0.077 | 0.033 | 1.080 | 1.011 | 1.153 | 2.13E-02 | 1.00E-05 | 4.50E-02 |
| C2 | T2DM | SIMEX | 28 | 0.099 | 0.037 | 1.104 | 1.027 | 1.186 | 1.23E-02 | 1.00E-05 | 4.21E-02 |

Please refer to Table 1 for abbreviations of the disorders and legends of Table 2.

**Table 4.**
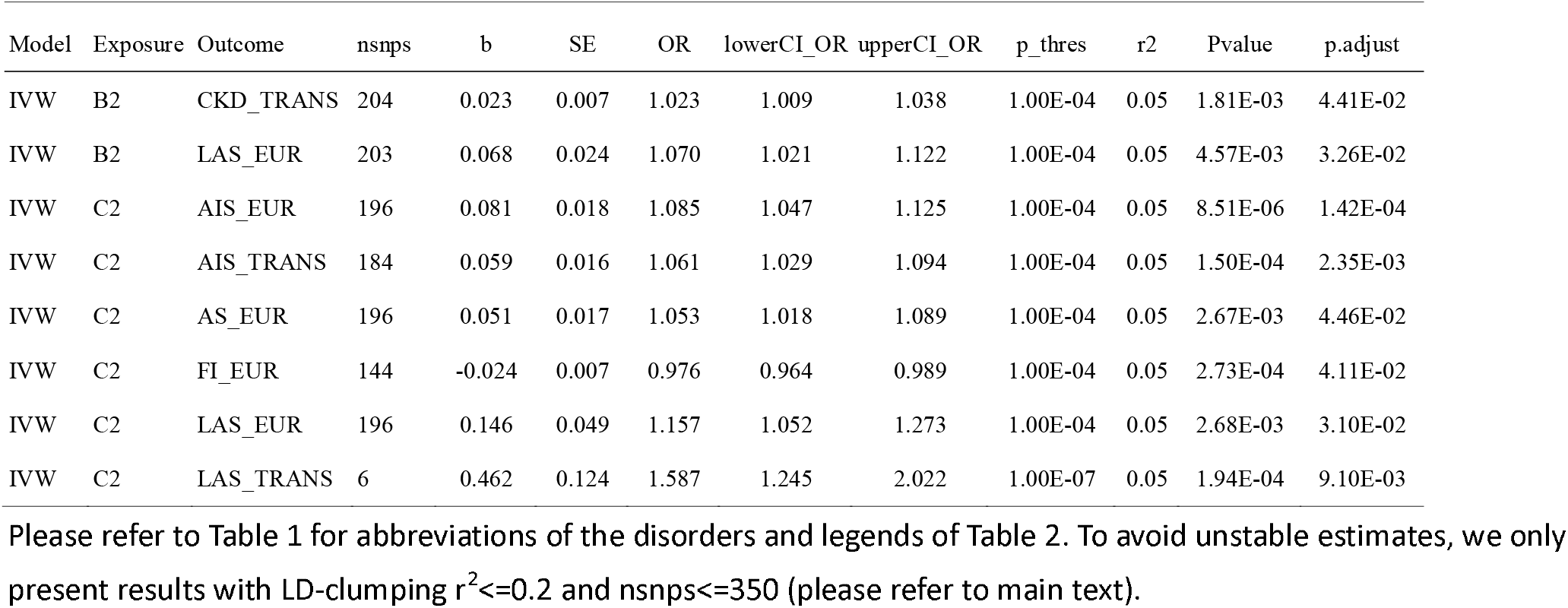
MR results with liability to COVID-19 as exposure and cardiometabolic disorders as outcome (correlated SNPs as instruments)

#### Independent SNPs analysis

In our primary analysis independent SNPs were used as instruments and MR was performed with each of the five methods across several pthres (Tables 2 and 3).

Across all CM disorders, *type 2 DM* (T2DM) showed highly consistent and significant causal relationships with COVID-19 in our analysis with independent SNPs (Table 3). We observed that liability to COVID-19 was causally linked to an increased risk of T2DM. This association was observed regardless of the type of infection (any COVID-19 infection[C2], hospitalized [B2] or critical illness[A2]), and across all five methods and multiple p-value thresholds. The effect size (coefficient) tends to be smaller with higher p-value thresholds, which may be the result of weak instruments bias and/or winner’s curse. Here we reported the effect size at the most stringent (lowest) p-value threshold since it is less subject to the above biases (same for other traits unless otherwise specified). For T2DM, with A2 (critical illness) as exposure, the OR per log-odds increase in exposure (roughly equivalent to 2.72-fold increase in the exposure risk) was 1.035 (CI: 1.006-1.065; pthres=5e-8; MR-IVW); with B2 (hospitalized infection) as exposure, the OR was 1.045, CI 1.009-1.081; pthres=1e-5; MR-IVW); with C2 (any infection) as outcome, the OR was 1.089 (CI 1.017-1.167 ; pthres=1e-5; MR-IVW). For the estimates by other methods and at other pthres, please refer to Table 3. Note that these figures are not directly comparable as the baseline risks for A2, B2 and C2 are not the same.

Besides T2DM, we also observed that liability to COVID-19 (hospitalized) infection may be causally associated with higher risk of chronic kidney disease (CKD) and several subtypes of stroke (Table 2). For CKD, the MR estimates were significant across multiple MR methods and for both European and trans-ethnic samples, when hospitalized infection (B2) was considered as exposure. The OR estimate was 1.030 based on MR-IVW (CI: 1.018-1.041; MR-pthres=1e-3) and slightly different with other methods (MR-Egger, OR=1.071; MR-RAPS, OR=1.034; weighted median[WM], OR=1.026; all at pthres=1e-3). For stroke, consistent associations were found for large artery stroke (LAS), mainly for B2 as exposure (MR-IVW, OR=1.057, CI 1.020-1.096; MR-RAPS, OR=1.063 CI 1.022-1.105; all at pthres=1e-3). The estimates from MR-Egger was larger (OR= 1.189, CI 1.073-1.316; pthres=1e-3). We also observed evidence of causal associations of hospitalized and general infection with cardioembolic stroke (CES) and small vessel stroke (SVS), but the significant results were restricted to the MR-Egger approach.

Several other results also passed FDR correction (FDR<0.05) and are briefly highlighted here. Hospitalized infection (B2) was observed to be causally linked to higher risk of heart failure (HF) across three MR methods. We also observed causal associations of any infection (C2) with lower fasting insulin and higher risk of atrial fibrillation (AF).

#### Correlated SNP analysis

The results of MR analysis with correlated SNPs are shown in Table 4 and Table S2. In general similar causal associations were observed with similar CM traits as above, using MR-IVW and GSMR. Note that to avoid instability of causal estimates, we restrict the number of SNPs and level of r^2^ so the results obtained here may not be fully comparable with those from independent SNP analysis.

We observed causal associations of A2 with T2DM and B2 with CKD and LAS, which were also observed in MR with independent instruments. We also observed causal links of C2 with higher risks of any stroke (AS) (MR-IVW, OR= 1.053, CI 1.018-1.089; pthres=1e-4; r^2^= 0.05), any ischaemic stroke (AIS) (MR-IVW, OR= 1.085, CI 1.047-1.125; pthres=1e-4; r^2^= 0.05) and LAS (MR-IVW, OR= 1.726, CI 1.166-2.553; pthres=5e-8; r^2^= 0.05), as well as lower fasting insulin (FI).

### MR results with COVID-19 as outcome and liability to cardiometabolic disorders as exposure

#### Independent SNPs analysis

Results are shown in Table 5 and S2. We observed that atrial fibrillation (AF) was causally associated with higher risks of critical (MR-IVW, OR= 1.098, CI 1.011-1.193; pthres=1e-5) and hospitalized infections. As for stroke, we observed that LAS was consistently associated with higher risks of critical, hospitalized or any infection (MR-IVW, OR= 1.019, CI 1.007-1.031; pthres=1e-4). Association with other types of stroke, including CES and SVS, were also observed but less consistent. For diabetes-related traits, T2DM was associated with higher risk of hospitalized infection, which was consistent across 3 MR approaches (IVW, MR-RAPS, GSMR) [MR-IVW, OR= 1.068, CI 1.019-1.119; pthres=1e-4]. Besides, we observed that T1DM (MR-IVW, OR= 1.027, CI 1.011-1.043; pthres=1e-2) and lower Insulin Sensitivity Index (ISI; adjusted for age and sex) were associated with higher probability of hospitalized infections [MR-IVW, beta= −0.090, CI (−0.145 - −0.036); pthres=1e-3]. Finally, we observed highly consistent causal associations of obesity with very severe, hospitalized or any infection across multiple MR approaches.

**Table 5.**
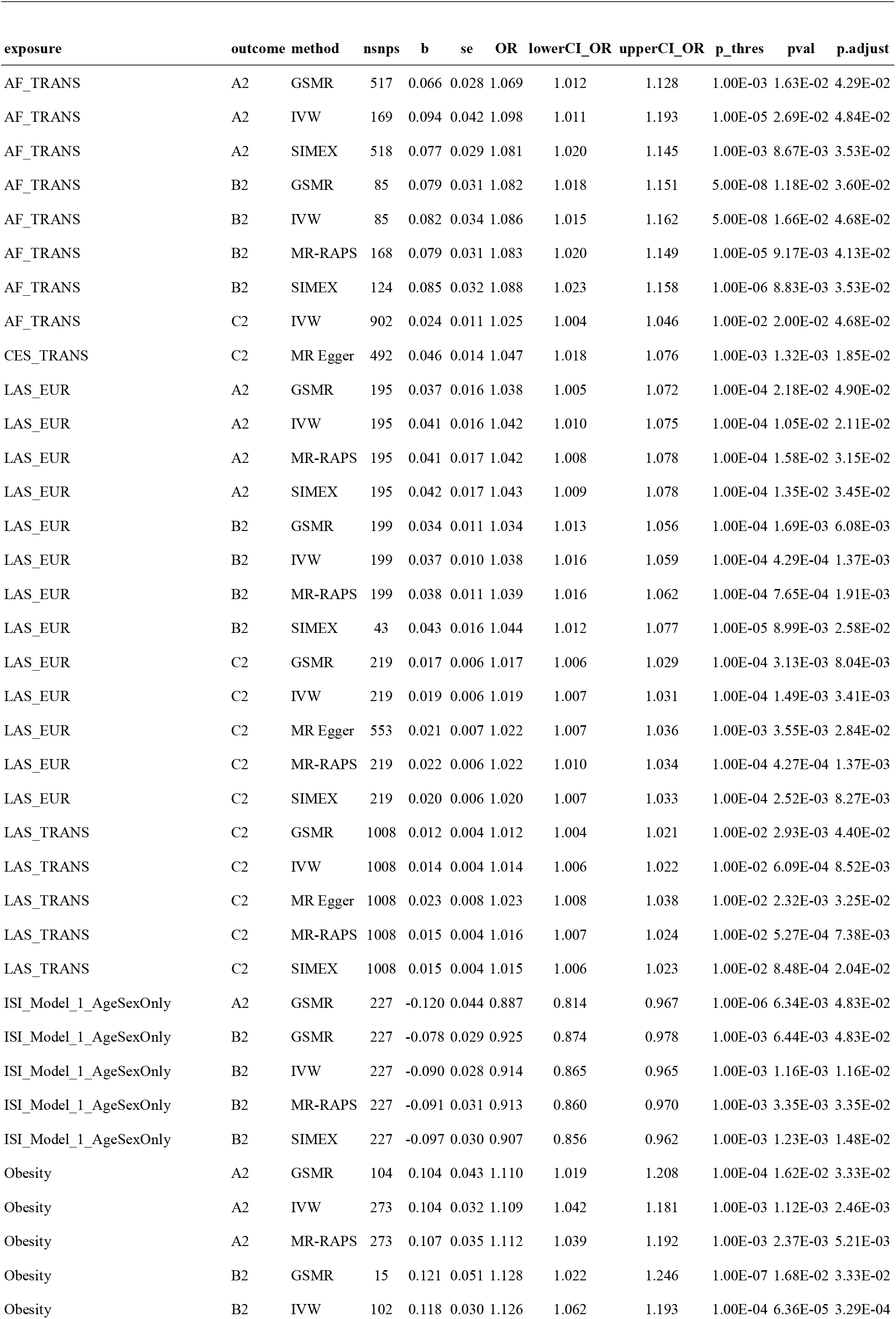
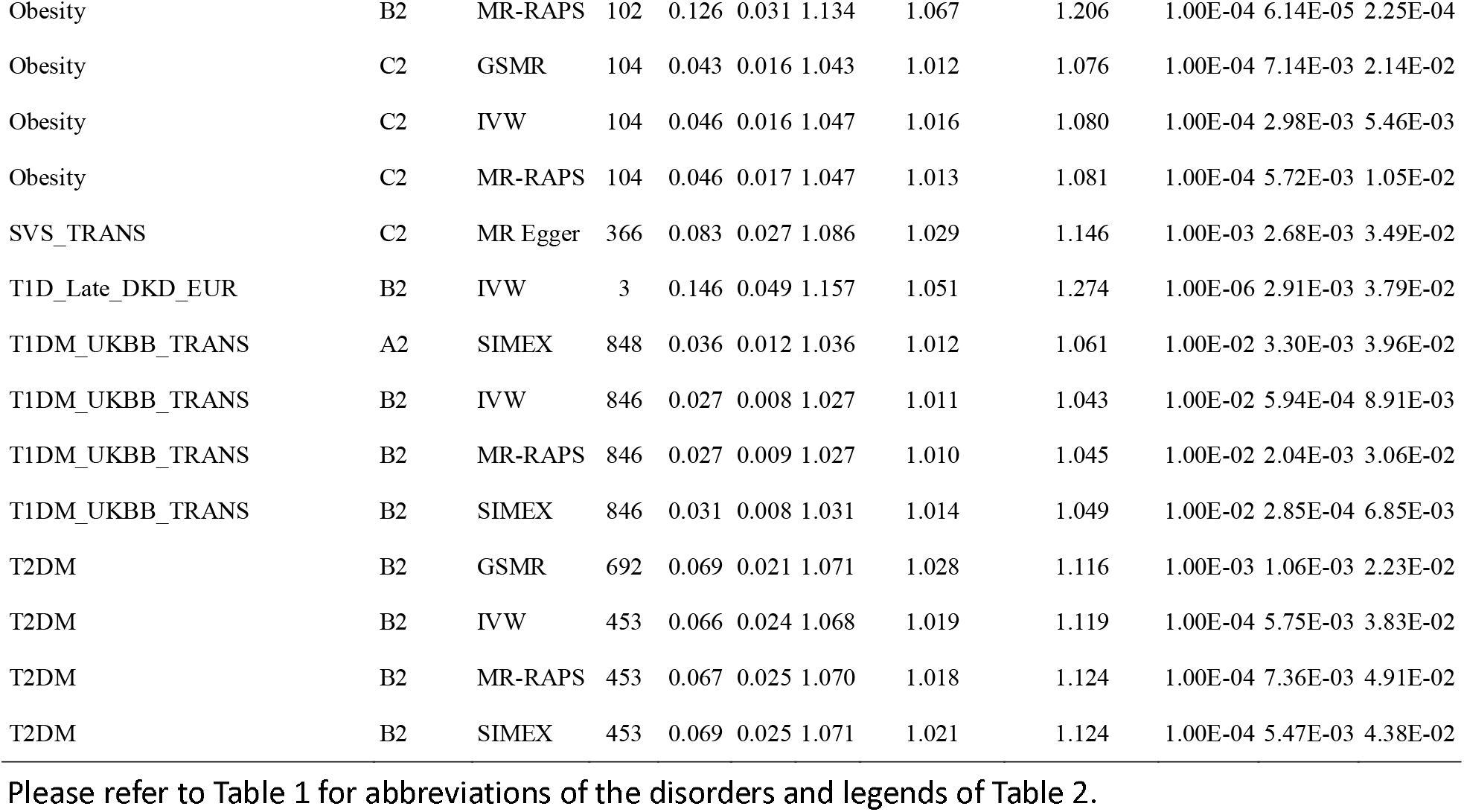
MR results with liability to COVID-19 as outcome and cardiometabolic disorders (CMD) as exposure (independent SNPs as instruments)

#### Correlated SNP analysis

The results are shown in Table 6 and S2. Similar to analysis with independent SNPs, we also observed significant causal associations of AF, stroke (LAS and any stroke), T1DM and obesity with any infection or severe infection. In addition, we also observed evidence of causal links of heart failure with infection/hospitalized infection. We also observed that early diabetic kidney disease (DKD) in T1DM and history of venous-thromboembolism were linked to higher susceptibility to infection and hospitalized disease. An inverse causal link between eGFR and severe or general infection was also observed.

**Table 6.**
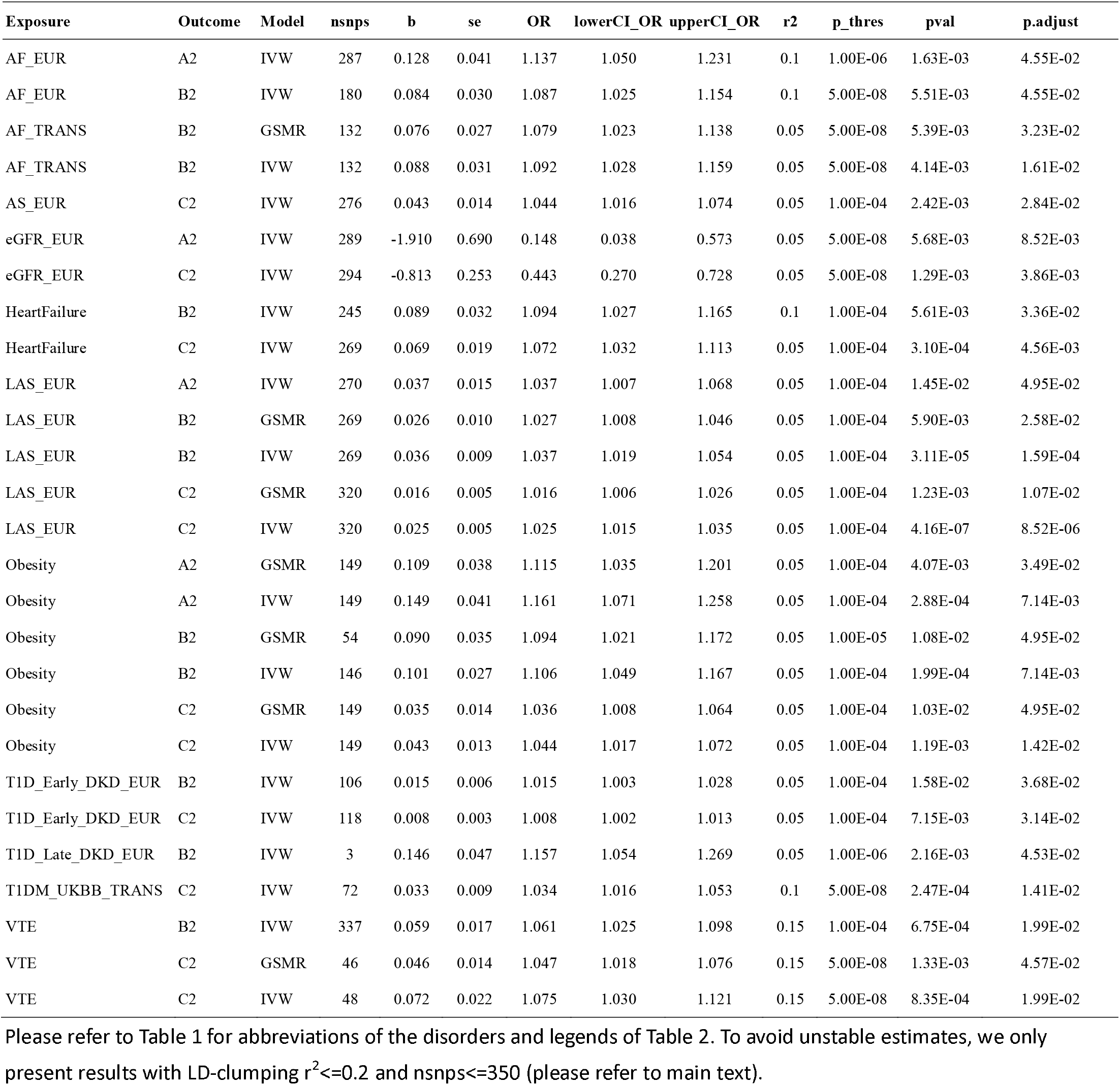
MR results with liability to COVID-19 as outcome and cardiometabolic disorders (CMD) as exposure (correlated SNPs as instruments)

#### Genetic correlation (rg) by LD score regression

Results which survive multiple testing correction (FDR<0.05) are shown in Table 7. Most of the CM traits implicated in MR analysis also showed significant genetic correlation. T2DM showed strongly statistical significant genetic correlation with both critical (rg=0.255, p=6.41e-7) and hospitalized infection (0.291, p=1.02e-7). CAD and obesity were also strongly genetically correlated with severe and hospitalized infection. Other disorders/traits showing significant and positive rg (FDR<0.05) included fasting insulin, CKD, gout, stroke (all stroke and any ischemic stroke) and urate levels. We note that all the significant results were restricted to A2 or B2 (critical or hospitalized disease) but not C2 (infection in general). Apart from the above, the intercept of rg analysis by LDSC can show the extent of sample overlap. We observed that intercept were mostly around zero (out of 116 pairs of traits with intercept estimates, only one has |intercept|>=0.02); the bias due to sample overlap is therefore likely small.

**Table 7.**
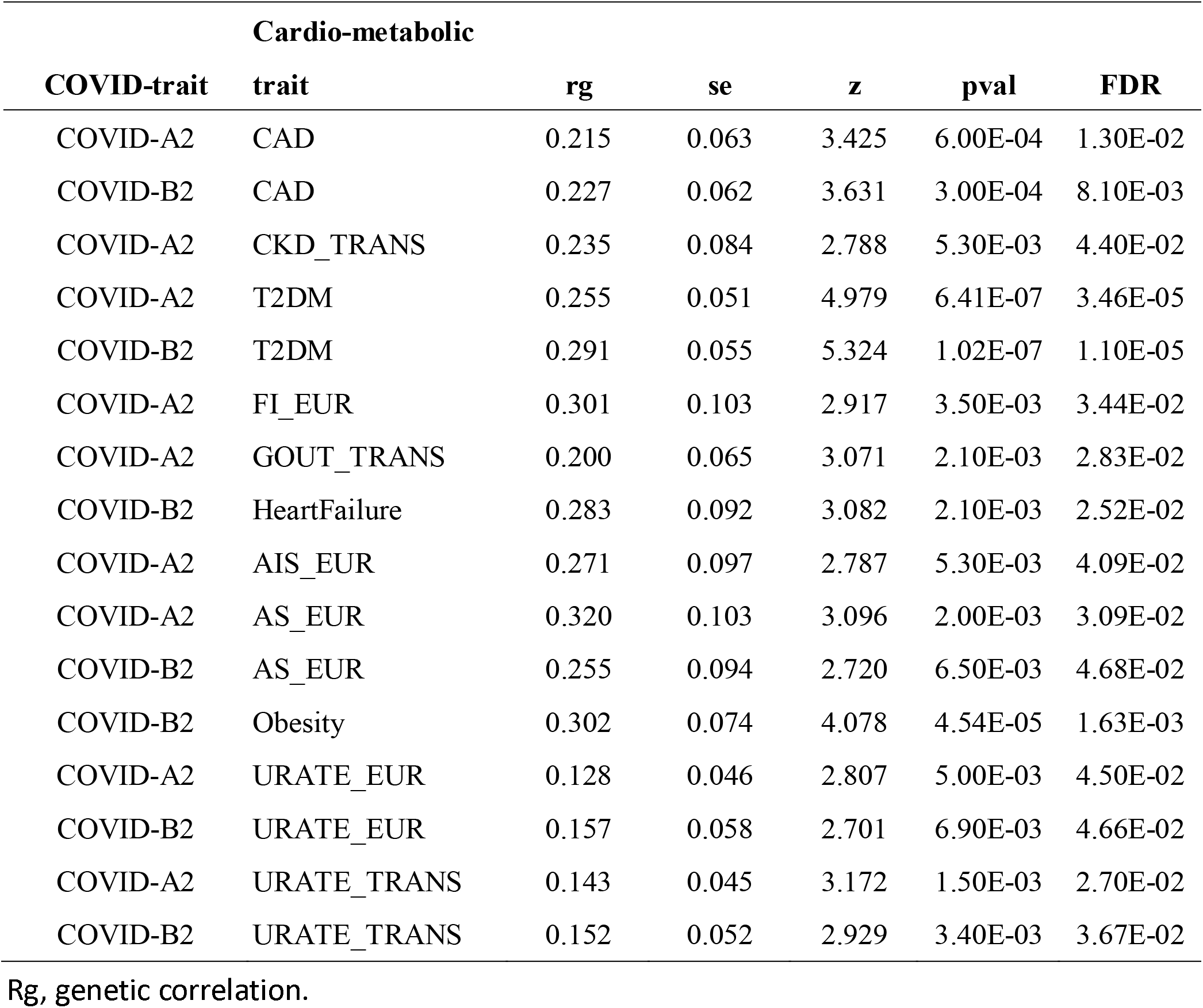
Genetic correlation analysis with LD score regression (results with FDR<0.05 are shown)

| **Cardio-metabolic** |  |  |  |  |  |  |
| --- | --- | --- | --- | --- | --- | --- |
| **COVID-trait** | **trait** | **rg** | **se** | **z** | **pval** | **FDR** |
| COVID-A2 | CAD | 0.215 | 0.063 | 3.425 | 6.00E-04 | 1.30E-02 |
| COVID-B2 | CAD | 0.227 | 0.062 | 3.631 | 3.00E-04 | 8.10E-03 |
| COVID-A2 | CKD_TRANS | 0.235 | 0.084 | 2.788 | 5.30E-03 | 4.40E-02 |
| COVID-A2 | T2DM | 0.255 | 0.051 | 4.979 | 6.41E-07 | 3.46E-05 |
| COVID-B2 | T2DM | 0.291 | 0.055 | 5.324 | 1.02E-07 | 1.10E-05 |
| COVID-A2 | FI_EUR | 0.301 | 0.103 | 2.917 | 3.50E-03 | 3.44E-02 |
| COVID-A2 | GOUT_TRANS | 0.200 | 0.065 | 3.071 | 2.10E-03 | 2.83E-02 |
| COVID-B2 | HeartFailure | 0.283 | 0.092 | 3.082 | 2.10E-03 | 2.52E-02 |
| COVID-A2 | AIS_EUR | 0.271 | 0.097 | 2.787 | 5.30E-03 | 4.09E-02 |
| COVID-A2 | AS_EUR | 0.320 | 0.103 | 3.096 | 2.00E-03 | 3.09E-02 |
| COVID-B2 | AS_EUR | 0.255 | 0.094 | 2.720 | 6.50E-03 | 4.68E-02 |
| COVID-B2 | Obesity | 0.302 | 0.074 | 4.078 | 4.54E-05 | 1.63E-03 |
| COVID-A2 | URATE_EUR | 0.128 | 0.046 | 2.807 | 5.00E-03 | 4.50E-02 |
| COVID-B2 | URATE_EUR | 0.157 | 0.058 | 2.701 | 6.90E-03 | 4.66E-02 |
| COVID-A2 | URATE_TRANS | 0.143 | 0.045 | 3.172 | 1.50E-03 | 2.70E-02 |
| COVID-B2 | URATE_TRANS | 0.152 | 0.052 | 2.929 | 3.40E-03 | 3.67E-02 |

Rg, genetic correlation.

## Discussion

### Overview

Here we have performed bi-directional MR analysis to uncover causal relationships between COVID-19 infection (and severe infection) with a wide range of cardiometabolic disorders. Overall we observed evidence that liability to COVID-19 or severe infection may be causally associated with higher risks of T2DM, CKD, stroke (especially LAS) and heart failure when compared to the general population. On the other hand, our MR results suggested that liability to AF, stroke (especially LAS), obesity, diabetes (T1DM and T2DM), insulin resistance and impaired renal function (low eGFR and DKD) may be causal risk factors for COVID-19 or severe disease.

This study has a number of strengths. A major novelty of this study is that we not only investigate causal risk factors for the infection, but also potential consequences as a result of COVID-19. The latter topic has been rarely addressed by causal inference methods such as MR. In addition, since COVID-19 is a new disease, many of its medium- or long-term consequences are difficult (or not possible) to be assessed by conventional observational studies. MR provides an alternative approach to uncover potential consequences of COVID-19 for further studies. We have employed the latest GWAS meta-analysis results from COVID-19 Host Genetics Initiative and a large variety of CM disorders and disease subtypes. We employed a variety of MR methodologies with strategies to improve statistical power, reducing the risk of false negative findings. At the same time, we also performed proper multiple testing correction (FDR) to control the expected proportion of false positives.

### Related MR studies

A few MR studies have been performed to investigate causal links between CM disorders and COVID-19. A very recent study ^22^ employed MR and found BMI as the only CM risk factor associated with higher probability of being tested positive and COVID-19 hospitalization, but the effect was non-significant after controlling for other CM traits including T2DM, CAD, stroke, and CKD. As discussed above, one main difference is that we also assess effects of COVID-19 on CM disorders. Here we used the latest release (release 5) of HGI instead of release 4 by Leong et al. ^22,24^, which contains a larger sample size. The CM disorders studied here were partially different from those by Leong et al., as we focused on diseases instead of general CM risk factors. We also covered a wider range of disease phenotypes such as stroke subtypes, AF, heart failure, DKD, among others. Consistent with Leong et al., we also found in this study that obesity is associated with higher risk of being tested positive as well as hospitalized or very severe infection. Here we also found that genetic liability to stroke (especially LAS) and T2DM were associated with being tested positive or severe infection, which were not reported in the previous work. The discrepancy between our study and Leong et al. may be due to different sets of summary statistics used (the latest release by HGI has large sample sizes and better power) and different MR methodologies used. Here we intend to improve statistical power by more liberal inclusion of SNPs as instruments. Also, we included stroke subtypes as phenotypes which may reduce sample heterogeneity and probability of detecting more specific associations. Another earlier report revealed that high BMI and smoking may be casually related to risk of COVID-19 and related hospitalization ^42^. Another study on lifestyle risk factors yielded a similar conclusion that genetically predicted BMI and smoking and also lower level of physical activities were associated with severe or hospitalized disease. ^43^. Yet another smaller-scale study (N∼1211) showed genetically predicted BMI and LDL were associated with infection risk ^44^.

### Relevance to other observational studies

Many of our findings are also supported by previous observational studies, which we shall highlight here. As for cardiometabolic consequences of COVID-19, as mentioned in the introduction, there has been reports of new-onset diabetes in COVID-19 and a recent meta-analysis showed a relatively high proportion of hospitalized patients (∼14.4%) had newly diagnosed diabetes. In addition, it was reported that COVID-19 may cause ketosis and ketoacidosis, and induced diabetic ketoacidosis in those with DM ^45^. There were also reports that COVID-19 may be linked to new-onset T1DM ^46,47^. Besides, in a recent Greek study, COVID-19 patients with and without diabetes showed admission hyperglycemia and those who were critically ill also showed reduced insulin secretion and poorer insulin sensitivity ^48^.

In this study we observed that liability to COVID-19 and hospitalized/critical infection were causally associated with T2DM. A number of mechanisms have been proposed for diabetogenic effect of COVID-19 (eg see ^17^). For instance, SAR-CoV-2 can attach to angiotensin-converting enzyme-2 (ACE2) receptors in beta cells of the pancreas, leading to acute impairment in insulin secretion. ^17^. An organoid study suggested that the virus can enter and damage the pancreatic beta cells ^49^. In addition, infection or severe infection may be associated insulin resistance ^50^. Conversely, both T1DM and T2DM have been shown to increase the morbidity and mortality from COVID-19 ^50-52^. Our study provides further support for a causal role of diabetes for severe infection.

There was also evidence that COVID-19 may be associated cardiac abnormalities and inflammation ^12^, and there were suggestions that COVID-19 can lead to worsening existing HF or new-onset HF ^53^. However, further prospective studies are required to establish association between the two entities and underlying mechanisms.

We also observed evidence that liability to (hospitalized) COVID-19 was causally associated with CKD. When comparing hospitalized patients with and without COVID-19, those with the infection were more likely to develop acute kidney injury (AKI) and require renal replacement therapy (RRT). AKI is an established risk factor for incident CKD ^54^. Another prospective study{Nugent, 2021 #74} reported that AKI patients with COVID-19, when compared to those without infection, had a steeper decline in renal function after discharge, even after controlling for comorbidities or severity of AKI. Conversely, we also found in our MR analysis (with correlated SNPs) that genetically predicted lower eGFR and T1DM-related DKD were associated with COVID-19. Observational studies ^55,56^ have shown a consistent link between renal failure and poorer prognosis and mortality from COVID-19.

As for stroke, our MR analysis suggested that liability to COVID-19 infection may be casually related to higher risks of ischemic stroke, especially LAS and possibly CES. On the reverse side, LAS was found to be associated with all COVID-19 phenotypes (A2, B2 and C2). Based on a recent systematic review by Fridman et al ^57^, stroke occurs at a relatively frequency (∼1.8%) among infected patients and carries a high mortality (34.4%). The risk of stroke was substantially higher (OR=7.6) when compared to patients with influenza, according to another study ^58^. Large vessel occlusion (LVO) was of high prevalence (46.9%) among all ischemic stroke patients with COVID-19, which is ∼1.5 times higher than the proportion of LVO in a population-based study (∼29.2%). The authors also found a very high prevalence of LVO (68.8%) in young patients (age<50) with relatively good past health (42.9% had no risk factors or related comorbidities), and proposed that hypercoagulability is a main cause of arterial thrombosis leading to stroke. Our finding suggested a bidirectional causal relationship between stroke (particularly LAS) and COVID-19 infection or severe infection, and is in line with findings from the above and other clinical studies. The possible mechanisms underlying the association between stroke and COVID-19 were reviewed elsewhere (eg ^59-61^).

We also found evidence that liability to AF was causally associated with critical illness or infection requiring hospitalization (A2/B2). Meta-analysis ^62^ showed that AF was associated with elevated risk of adverse outcome among COVID-19 patients, including mortality. Another study ^63^ also showed decreased survival in patients with AF. On the other hand, AF may also be a consequence of COVID-19 infection ^64,65^. From our MR analysis, we did find liability to infection (C2) was associated with increased AF risk (OR=1.100, CI 1.040 −1.163, pval=8.93E-04), supported by the GSMR approach, although we did not observe consistent significant (FDR<0.05) associations across other methods or thresholds. Besides, it is also intriguing to note that we found tentative evidence that the infection may be associated with cardio-embolic stroke (CES), mainly supported by MR-Egger analysis. AF is a known and strong risk factor for CES ^66^ and may represent one of the mechanisms of increased stroke incidence in infected patients.

### Limitations

There are several limitations for this study. We have employed the latest and largest GWAS summary statistics to date for COVID-19, however the data is based on meta-analysis of a large number of separate studies, and the samples may be heterogeneous. For example, the baseline clinical features, demographics, comorbid disease patterns etc. of patients or controls may differ across cohorts. The control population were unscreened, and therefore asymptomatic patients or those with mild symptoms may be missed. Hospitalization is in general a reasonably good proxy for moderate or severe illness, but the criteria for hospitalization may still differ across countries and cohorts. Although the sample size of COVID-19 GWAS is already quite large, the number of critically ill and hospitalized cases may still be relatively limited when compared to the current standards. As such, a lack of association may be due to lack of power. The same also applies to GWAS of CM disorders or traits with smaller sample sizes.

MR is a highly useful methodology that has been successfully applied in cardiovascular medicine ^67^ and other fields to evaluate causal relationships. However, it is not without limitations. One concern of MR is horizontal pleiotropy (an instrument associated with the outcome not through the exposure), which we have tried to address with different methodologies of different principles. However, each method has its own assumptions (e.g. InSIDE assumption for MR-Egger, systematic and idiosyncratic pleiotropy for MR-RAPS, >50% valid instruments for WM etc.), it is not possible to know a priori the pattern of pleiotropy and whether assumptions are fully fulfilled. However, many of the associations reported here were relatively robust across multiple MR methods. We also note that for binary exposures, the interpretation of MR may not be very intuitive ^37^. The effect estimate cannot be directly interpreted as the OR of developing the outcome when the exposure (e.g. a disease) is present, but may be conceptualized as the effect per unit (log-odds) increase in the *liability* to exposure [roughly, every 2.72 times increase in Pr(exposure)]. For rare exposures (e.g. A2), this may reflect an increase in prevalence from e.g. 0.3% to 0.82%, and one may not expect this effect to be large. As such, although the ORs reported in this study were mostly modest, they should be interpreted with the above caveats in mind. Moreover, even if the effect sizes (OR) are modest (e.g. 3% increased risk of T2DM due to the infection), given the very large number of people affected by COVID-19 to date, the absolute number of people affected and hence public health burden may still be substantial. Another limitation is that the MR analysis cannot tell if the potential consequences of COVID-19 will likely occur in the short- or long term; it only suggests a causal relationship.

As explained earlier, based on properties of MR and various simulation studies (by us and others), we believe the significant MR results are unlikely to be explained by inflated type I error rate, but the effect size estimates may be biased towards zero so should be interpreted carefully. In addition to measurement error of exposures which may lead to conservative causal estimates, it has been raised that selection bias (winners’ curse) may bias estimates of SNP-exposure coefficients away from zero. Previous studies suggested that this bias is actually present in all MR studies, even if a stringent p-value threshold is used. However, inclusion of a larger number SNPs may exaggerate the problem. This bias is again conservative in two-sample MR. Intuitively, the wald ratio estimate is SNP-outcome coefficient divided by SNP-exposure coefficient; if the latter (i.e. denominator) is biased away from zero, the causal estimate will be biased towards the null ^68^.

## Conclusions

To our knowledge, this is the most comprehensive MR study to date investigating ***bi****-directional* causal links between COVID-19 and CMD. In summary, this study provides evidence for tentative causal relationships between liability to (severe) COVID-19 infection and a number of CM disorders. We found that the infection may be associated with higher risks of several CM disorders such as T2DM and stroke, while many CM disorders may be causal risk factors for infection or severe disease. Due to the limitations described above, these findings should not be regarded as confirmatory and needs to be further replicated in larger studies and by prospective clinical studies. The underlying mechanisms also require further investigations.

## Data Availability

All GWAS summary statistics were downloadable from publicly available sources. Results of the current analyses were available in the main paper and supplementary tables.

## Acknowledgements

This work was supported partially by a National Natural Science Foundation China (NSFC) grant (81971706), Lo Kwee Seong Biomedical Research Fund from The Chinese University of Hong Kong (CUHK) and a Direct Grant from CUHK. We thank Mr. Kenneth C.Y. Wong for preparation of some GWAS summary datasets.

## Author Contribution

Conception and design: HCS. Analytic methodology: HCS (lead) and XY. Data analysis: XY (main) and HCS (main), with input from CKLC, XY and JQ. Data interpretation: HCS, XY, CKLC, JQ, SR. Supervision of study: HCS. Drafting of manuscript: HCS, with input from XY.

## Supplementary Information

All supplementary Tables and notes are available at the journal’s website and at https://drive.google.com/drive/folders/17DoiLSjL_tNyJTaOXnKdcJM5VDAe6GUL?usp=sharing

## Conflicts of interest

The authors declare no conflict of interest.

## Notes

### Competing Interest Statement

The authors have declared no competing interest.

### Author Declarations

Only GWAS summary statistics were used in this study without individual-level data. The respective studies have obtained proper IRB approval.

